# Outcome Of Total Surgical Debridement Of Covid Associated Skull base Mucormycosis Based on a New Surgical Staging System: Evidence From A Cohort Study

**DOI:** 10.1101/2022.11.04.22281828

**Authors:** Lekshmy R Kurup, Harshita Singh, Shilpee Bhatia Sharma, Puya Dehgani-Mobaraki, Asiya Kamber Zaidi, Narayanan Janakiram

## Abstract

**Purpose:** To propose a surgical staging system with management protocol for post-covid Rhino-Orbito-Cerebral Mucormycosis (ROCM) with central skull base osteomyelitis.

**Methods:** A prospective cohort study of post-covid ROCM patients between May 2021 and January 2022. Patients were assessed radiologically and staged from I to V. Follow up period was 6 to 18 months and the surgical outcome was assessed.

**Results:** Total of 193 patients (129 primary and 64 revision). Maxilla was found to be the epicenter of anterior disease (69.3%) and pterygoid wedge, the epicenter of posterior disease (85.6%). More than 65% of our patients, at the time of presentation, presented with ROCM with involvement of the central skull base. Intracranial disease was noted in 13.9% of patients and the mortality rate was 6.2 %.

**Conclusion:** This staging system provides a systematic step-by-step protocol for the management of ROCM, with emphasis on meticulous disease clearance at the central skull base.

## INTRODUCTION

Coronavirus disease 2019 (COVID-19) has been associated with multiple opportunistic bacterial and fungal infections. Covid-19 Associated Mucormycosis has been reported in many countries like Austria, Brazil, France, India, Iran, and the US. A steady increase in CAM in the Indian context appears to be due to the large diabetic population and imprudent use of corticosteroids by the healthcare workers. Other well-known causes for Mucormycosis include; neutropenia, organ or bone marrow transplantation, burns, hematologic malignancies, post chemotherapy status, and dialysis. This rare, intriguing entity became of immense public health importance due to its angio invasive nature leading to high morbidity and fatality rate.^(1)^

The complex pathogenesis of Rhino-Orbito-Cerebral Mucormycosis(ROCM)may be attributed to hypoxia, hyperglycemia (diabetes, steroid-induced), acidic medium (metabolic acidosis, diabetic ketoacidosis), high iron levels (increased ferritin), and decreased phagocytic activity of WBC due to immunosuppression along with prolonged hospitalization with or without mechanical ventilation.^(2)^ Covid 19 associated ROCM is the most frequent presentation of the disease. Given its proximity to the central skull base, the infection travels through fissures and suture lines to the Haversian system of the compact bone. Further angioinvasion and arterial thrombosis lead to necrosis of the tissues.

A high index of suspicion and accurate assessment of predisposing factors is the cardinal prerequisite for diagnosis of Mucormycosis. Diagnostic nasal endoscopy, imaging studies and pathological confirmation contribute to diagnosis and surveillance. MRI is essential for determining the overall extent of the disease, marrow and extraosseous soft tissue involvement and intracranial extension.

There was a steep increase in the number of Mucormycosis cases and since the response and tolerability of the optimal dose and duration of therapy in ROCM associated Central Skull Base Osteomyelitis (CSBO) were unclear and unguided, it prompted us to come up with an evidence-based protocol for ROCM.

These lacunae prompted us to conduct a review of our own consecutive patients with ROCM and CSBO, to know its temporal association with comorbidities and the overall characteristics of patients with its outcome. It further describes surgical strategies of aggressive debridement with optimal medical management to curtail fatality. The present study is the first attempt in the literature to propose a clinical-surgical management algorithm to elucidate disease patterns and multidisciplinary treatment strategies including skull base debridement for ROCM with CSBO.

## MATERIALS AND METHODS

We performed a prospective cohort study of 193 adult patients who presented at our tertiary referral center between May 2021 and January 2022 with clinically and microbiologically proven ROCM and history of recent SARS-CoV-2 infection (delta variant) or COVID-19 sequalae. The demographics, predisposing factors, symptoms, radiological, laboratory, histological and microbiological data were collected with the patient’s consent. The authors assert that all procedures contributing to this work comply with the ethical standards of the national and institutional guidelines on human experimentation and with the Helsinki Declaration of 1975, as revised in 2008.

A total of 129 primary cases were included in this cohort study with 64 previously operated cases. Non-covid ROCM with CSBO were excluded from the study. The follow-up period ranged from 6 to 18 months post-surgery.

The diagnosis of COVID-19 was based on any one of the following: a positive result on Reverse Transcriptase-Polymerase Chain Reaction (RT-PCR) test on nasopharyngeal or oropharyngeal swabs, a positive result on rapid antigen test (RAT) and in clinically symptomatic cases, a computed tomography (CT) chest scores followed by positive RT-PCR/RAT. Cases were categorized as proven ROCM if clinical, radiological, microbiological or pathological features were positive.

Patients were defined as recovered from covid infection if the repeat RT-PCR test was negative or two weeks had elapsed since the diagnosis.

Magnetic resonance imaging (MRI) of Paranasal sinuses with Orbit and Brain, and (T1W, T1W STIR (short tau inversion recovery) with contrast, T2W, T2W FS and Diffusion Weighted Imaging (DWI)) with Computed Tomography of Paranasal Sinus (CT PNS) was performed to assess the extent of the disease. Patients with clinical features suggestive of palatal involvement were evaluated using Cone Beam Computed Tomography Scan (CB-CT) of maxilla. In cases with evidence of gross erosion of bone or frontal table involvement, an MDCT with 3-D facial reconstruction was performed. A KOH smear and tissue biopsy was done for all patients pre-operatively.

A multidisciplinary team was formed, which comprised of otorhinolaryngologists, radiologists, maxillofacial surgeon, ophthalmologist, nephrologist, endocrinologist, neurosurgeon, critical care physician and anesthesiologist. Glycemic control and management of co-morbidities were strictly monitored and adjusted accordingly. The nutritional status was considered crucial, and a high protein diet with supplements was assured.

The advances in understanding complex endoscopic skull base anatomy and instrumentation have evolved multiple approaches to eradicate central skull base osteomyelitis and improve clinical outcomes. After a meticulous analysis of the disease extent, a tailored, suitable endoscopic or combined approach was selected. A validated classification system would map the extent of the disease, which would help the surgeon to follow the proposed surgical steps and principles. This staging system can standardize treatment protocol and could be utilized for disease prognosis and prediction of complications. Extensive debridement was required to remove infected necrotic tissue to provide clean, viable margins.

## PRE-OPERATIVE SURGICAL PLANNING

All patients were pre-operatively assessed by the anaesthetist for airway, co-morbidities, blood parameters and treatment history. Pre-medications were given prior to surgery for reducing anxiety and facilitating induction. Benzodiazepines and H2 blockers were prescribed the night before as well as in the morning of surgery. If the systolic blood pressure (BP) of patient was more than 100 mm of Hg, Beta blockers were administered to ease hypotensive anaesthesia. Every patient and his relatives were explained in detail about the procedure and its possible complications.

All the involved tissues and surrounding tissues were sent for histopathology to assess the degree of involvement and confirm the diagnosis. In revision cases on antifungal therapy, PCR-based molecular techniques have been shown to have high sensitivity for confirming fungal strain.

The vascularity and mucosalization were assessed endoscopically every week. In those patients with Superior orbital fissure syndrome, ptosis and restricted eye motility had partially resolved during the follow-up, but visual loss persisted. The treatment outcome was evaluated with serial MRI scans after one month, third month, sixth month and one year after surgery.

## RESULTS AND ANALYSIS

The mean age was 48.9 years (10.6). Out of the total 193 patients, 154 (79.8%) were males and 39 (20.2%) were females. A total of 129 cases (66.8%) accounted for primary and 64 cases (33.2%) for revision cases. Associated Co-morbidities were also considered and documented. Details are depicted in Figure 1.

**Fig 1.**
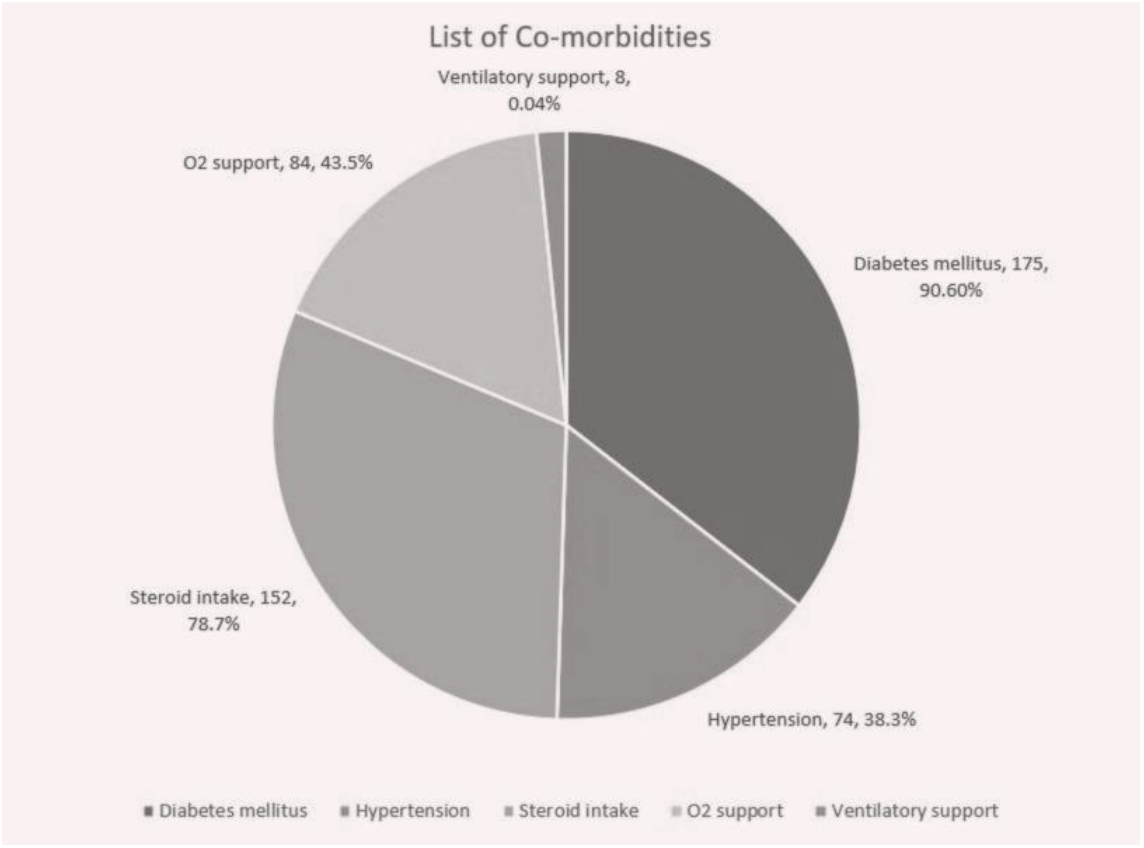

Stage wise radiological distribution of primary and revision cases as per JSPM staging is depicted in fig 2.

**Fig 2.**
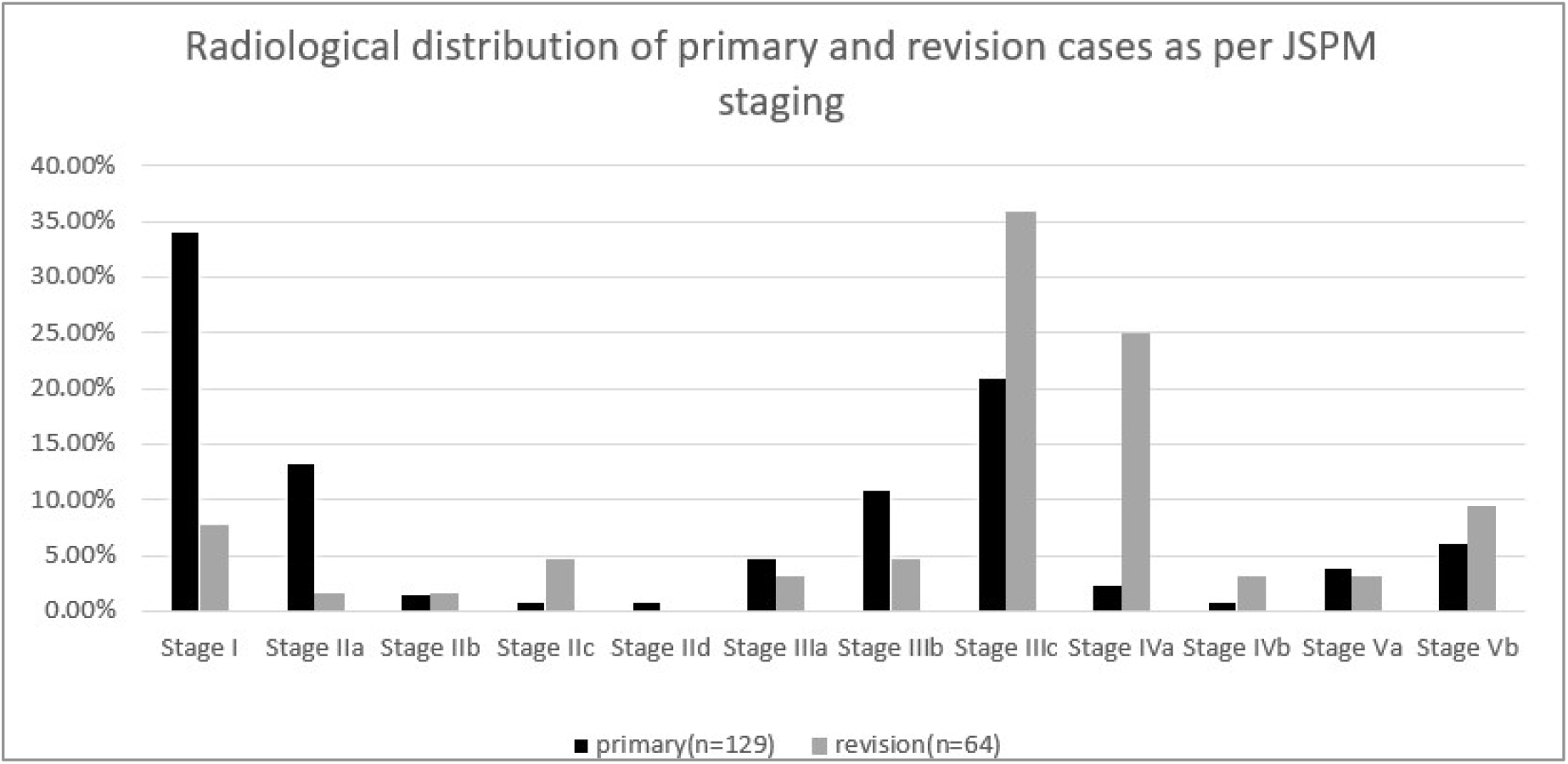

In primary disease stage I (34.1%) was most common followed by stage IIIC (20.9%). In revision cases stage IIIC (35.9%) numbered the most followed by stage IVA (25.0%).

Anterior disease is from stage I to stage IID and posterior disease from stage IIIA to VB. Out of the total 193 cases, 38.8% (n=75) patients were staged as anterior disease and 61.2% (n=118) patients were staged with posterior disease.

Involvement of maxilla was found to be 69.3% (n=52) in anterior disease(n=75) compared to 51.7% (n=61) in posterior disease (n=118) (p value = 0.015 statistically significant), thus supporting our hypothesis that maxilla forms the epicentre in anterior disease.

Similarly, out of 118 patients with posterior disease, n=101 patients (85.6%) showed involvement of pterygoid wedge as compared to n=3 patients (4%) among the anterior disease (n= 75, P value < 0.001). This again substantiates our hypothesis that pterygoid wedge is the epicentre in posterior disease.

Most common site among the primary 129 cases was maxilla (66%) and least common site was orbital apex (3.1%). Among the 64 revision cases, most common site was pterygoid wedge (75%) and least common was cavernous carotid and skin (both 3.1%). Skin involvement was only seen in 4 patients out of which three were minimal involvement.

The JSPM classification was applied to intra-operative findings too, the Intra-op staging is depicted in Figure 3:

**Fig 3.**
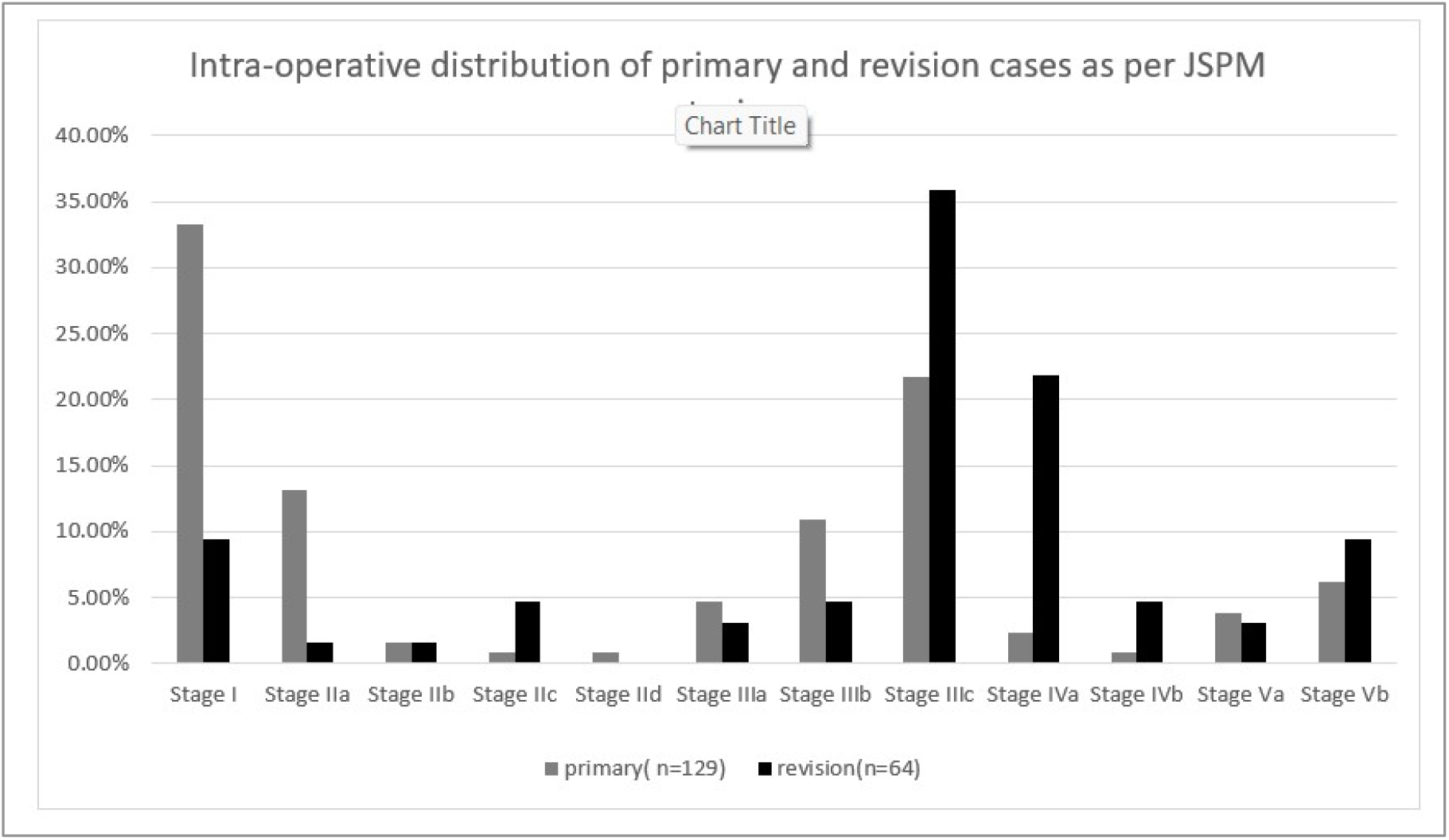

Intra-operative staging correlated with radiological staging in disease distribution except in five patients.

Comparing the site distribution of disease in primary and revision disease and their statistical significance is elaborated in table 2.

**Table 1:**
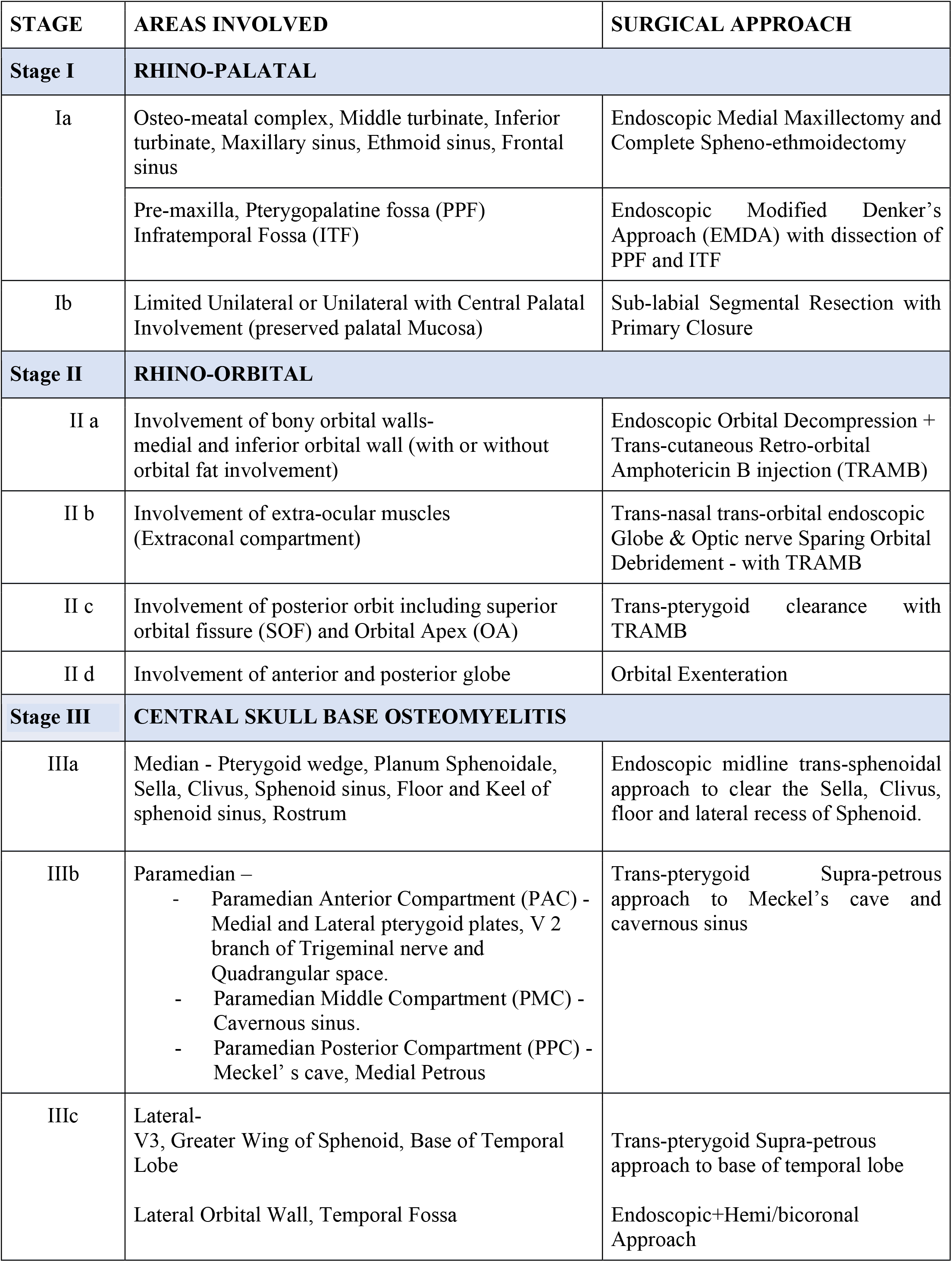
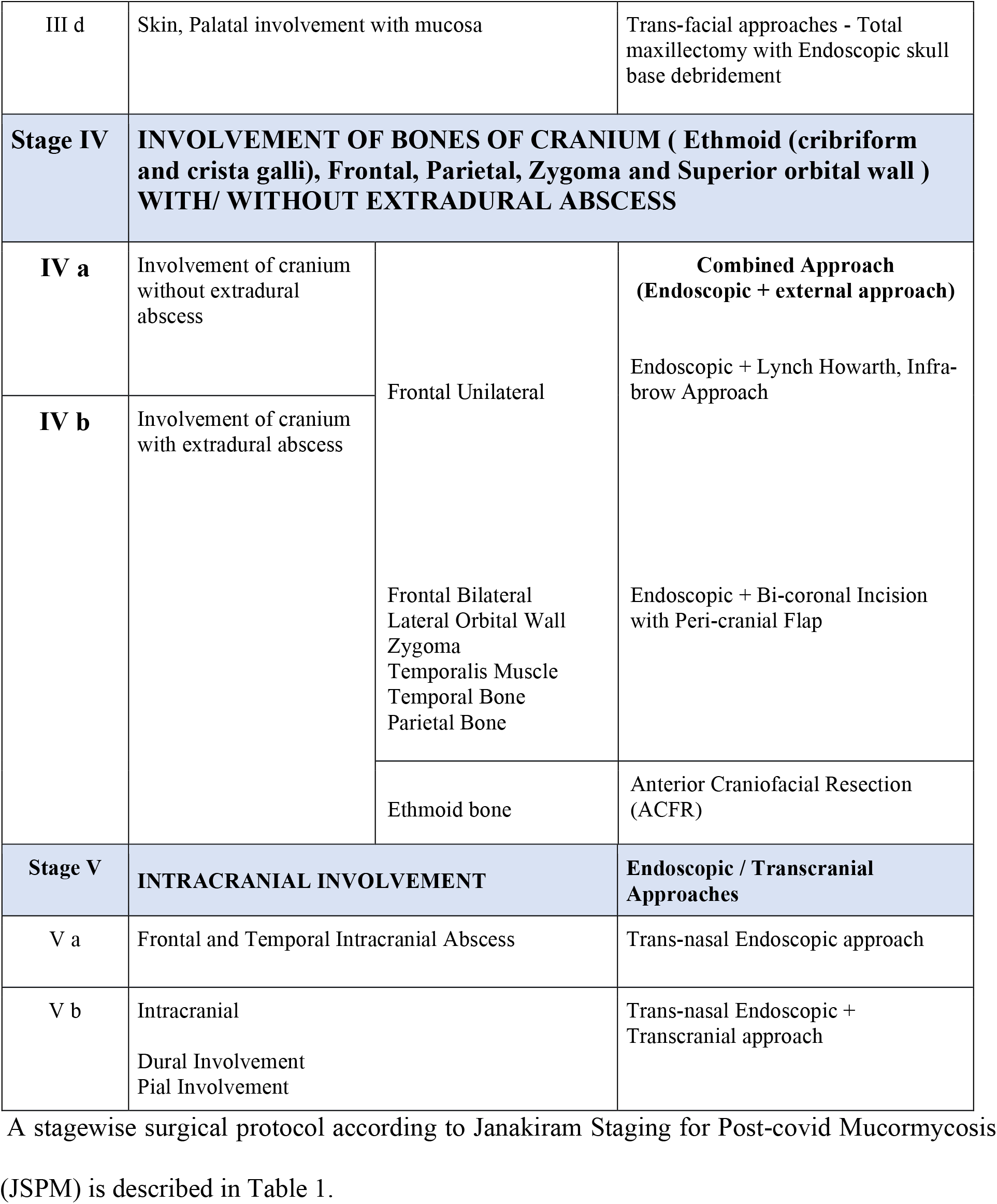
JSPM Staging system with Surgical protocol

**Table 2:**
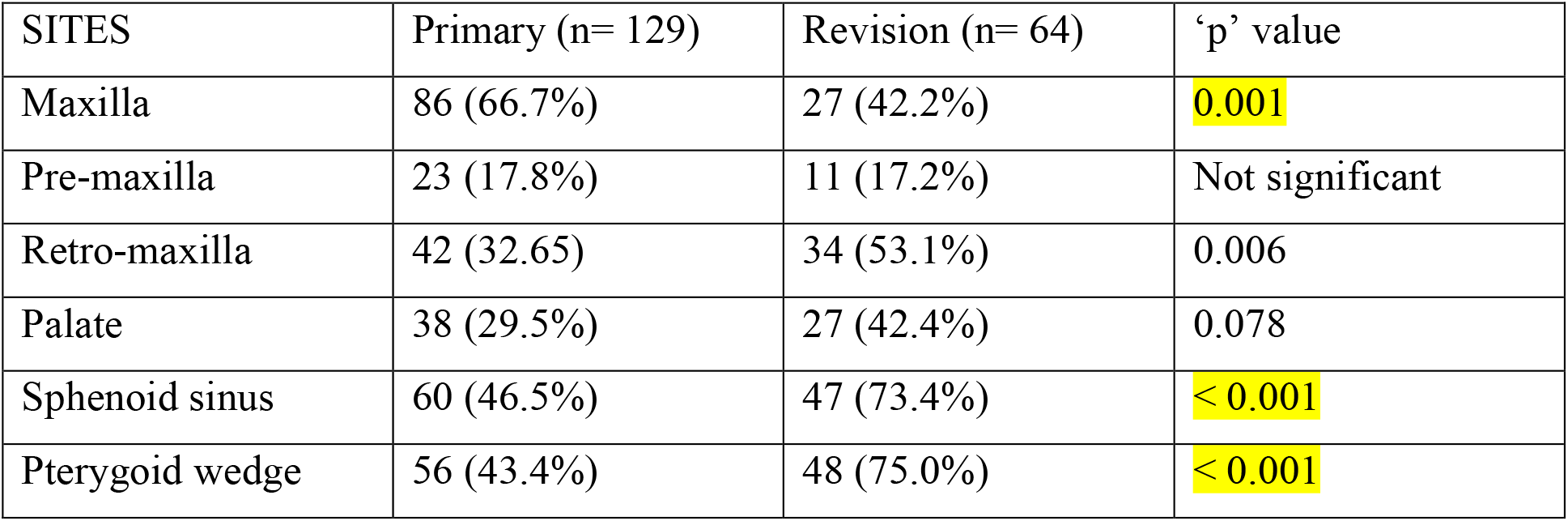
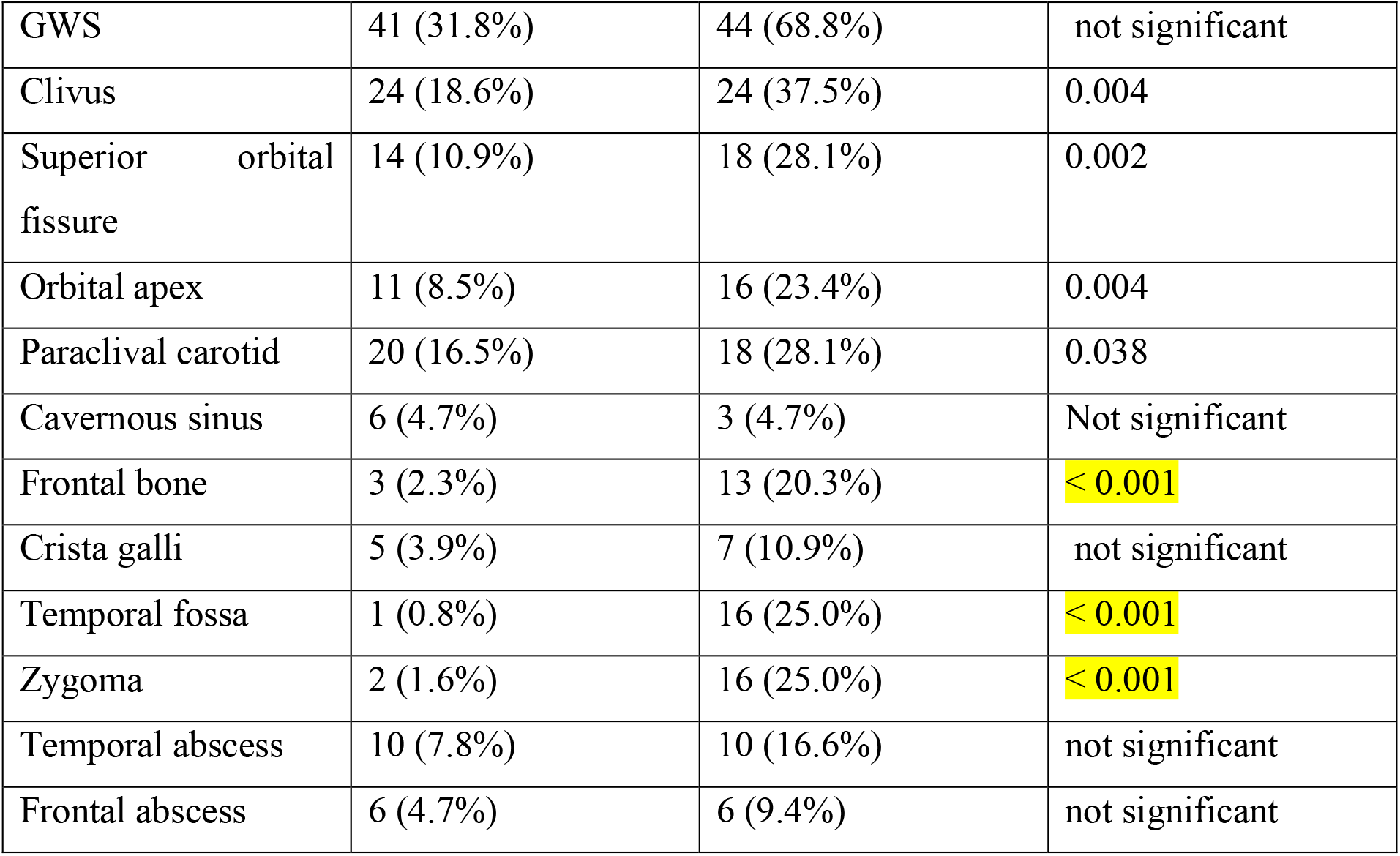

It was observed that maxilla was significantly more involved in primary disease than revision cases.

The retro-maxilla was involved in 53.1% of revision cases and 32.6% of primary cases (p value< 0.006). Subsequently the pterygoid wedge was also involved in 75% of revision cases and 43.4% of primary cases. This substantiates that the spread of disease to posterior skull base is faster from the retro-maxilla.

Out of the n=75 patients with anterior disease, n=65 patients were primary and n=10 were revision cases. The frequency of involvement of sites is given in table 3. Maxilla is involved in 73.8% of primary cases whereas only 40% of revision cases showed involvement of maxilla. This is possibly because they have already been operated and maxillary clearance was given previously.

**Table 3:** Anterior disease-sites involved in primary and revision cases

| Site | Primary (n=65) | Revision (n=10) |
| --- | --- | --- |
| Pre-maxilla | 13 (20.0%) | 1 (10%) |
| Maxilla | 48 (73.8%) | 4 (40.0%) |
| Anterior Ethmoid | 35 (53.8%) | 0 (0.0%) |
| Retro-maxilla | 25 (38.5%) | 1 (10.0%) |
| Frontal sinus | 17 (26.2%) | 3 (30.0%) |
| Palate | 20 (30.8%) | 4 (40.0%) |

Similarly, out of the 118 patients with a posterior disease n=64 were primary and n=54 were revision cases. The different sites involved and their frequency is given in table 4. In primary and revision cases, involvement of pterygoid wedge and sphenoid was significant.

**Table 4:** Posterior disease-sites involved in primary and revision cases

| Site | Primary (n=64) | Revision (n=54) |
| --- | --- | --- |
| Sphenoid | 57 (89.1%) | 47 (87.0%) |
| Pterygoid Wedge | 53 (82.8%) | 48 (88.9%) |
| Greater wing of Sphenoid | 40 (62.5%) | 44 (81.5%) |
| Clivus | 24 (37.5%) | 24 (44.4%) |
| Paraclival carotid | 20 (31.2%) | 18 (33.3%) |
| Cavernous Sinus | 6 (9.4%) | 3 (5.6%) |
| Frontal Bone | 3 (4.7%) | 13 (24.1%) |
| Crista Galli | 4 (6.2%) | 7 (13.0%) |
| Temporal Fossa | 0 (0.0%) | 16 (29.6%) |
| Zygoma | 1(1.6%) | 16(29.6%) |
| Temporal Abscess | 7 (10.9%) | 9 (16.7%) |
| Frontal Abscess | 5 (7.8%) | 6 (11.1%) |
| Cavernous carotid | 3 (4.7%) | 2(3.7%) |
| Petrous carotid | 2 (3.1%) | 3 (5.6%) |

Figure 4 depicts the primary and revision cases based on site of involvement (bilateral disease involvement in the frontal sinus, sphenoid sinus, pterygoid wedge, greater wing of sphenoid and palate for n=193 cases). Bilateral involvement was not seen in orbit, superior orbital fissure, orbital apex or intracranial abscess.

**Fig 4.**
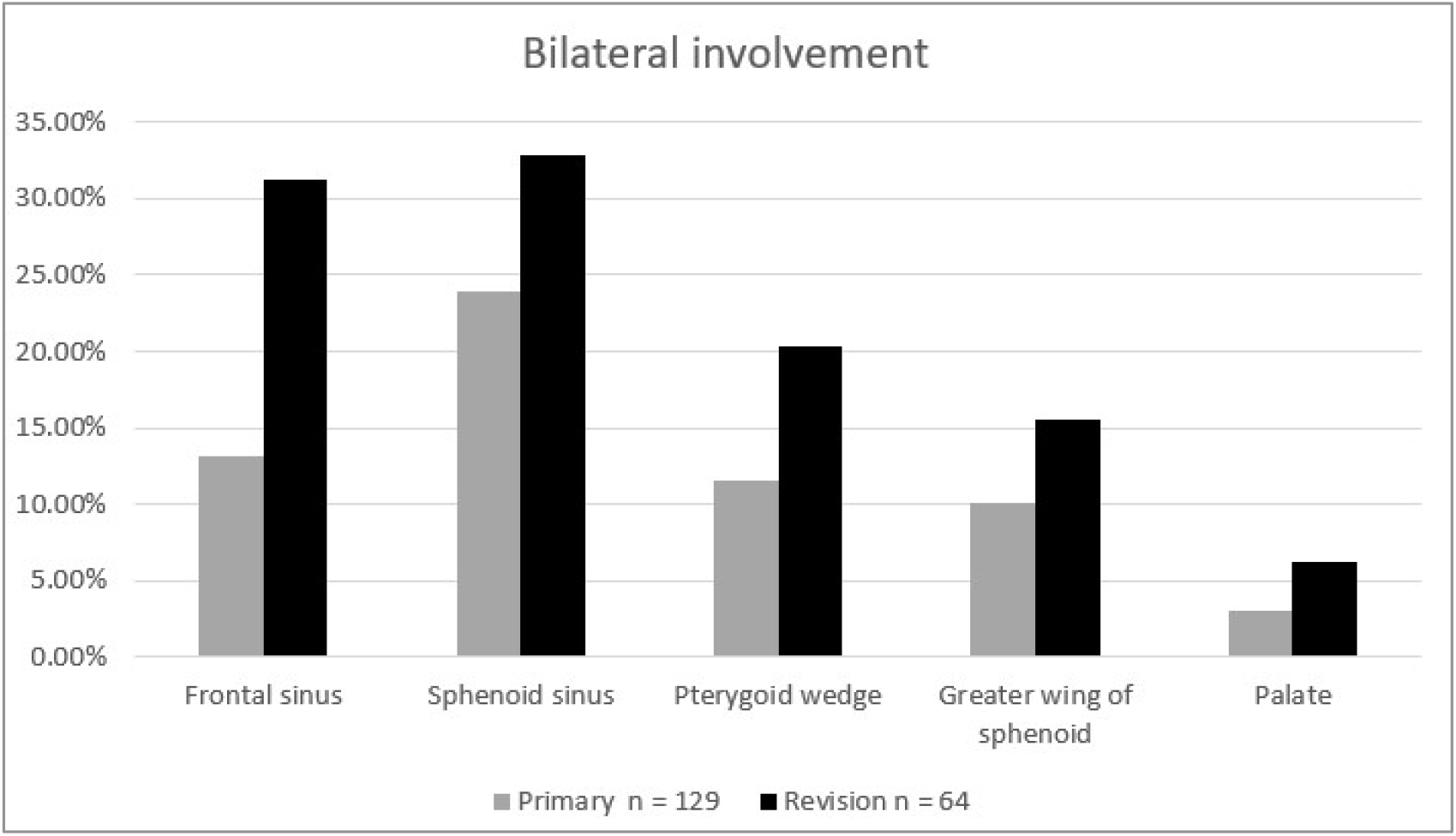

Table 5 depicts the involvement of orbit in primary and revision cases for n=193 cases.

**Table 5:**

| Orbital wall | Primary (n = 129) | Revision (n = 64) | p value |
| --- | --- | --- | --- |
| Medial wall | 18 (14%) | 19 (29.7%) | 0.01 |
| Inferior wall | 16 (12.4%) | 18 (28.1%) | 0.007 |
| Superior wall | 1 (0.8%) | 4 (6.2%) | 0.024 |
| Lateral wall | 1 (0.8%) | 4 (6.2%) | 0.024 |
| Orbital exenteration | 1 (0.8%) | 11 (17.2%) | < 0.05 |

With respect to orbital disease, individual wall involvement as well as number of exenterations were more in revision cases.

### Residual disease

In the entire cohort of 129 primary cases, n=7 patients had disease residue. Among the 64 revision cases, only n=2 cases recurred, both patients were operated for temporal lobe abscess and subsequently underwent craniotomy and abscess drainage. Currently they are on follow up and disease free.

Details of possible cause for residue and the time interval between surgery and residual disease is stated in table 6.

**Table 6:**

| Serial no | Primary stage | Revision stage | Cause for residue | Time Interval |
| --- | --- | --- | --- | --- |
| 1 | I | I | Amphotericin intolerance | 3 months |
| 2 | I | I | Palatal residue | 3 months |
| 3 | I | I | Amphotericin intolerance | 4 months |
| 4 | IIIB | IIIC | Amphotericin intolerance | 4 months |
| 5 | IIIB | IIIC | Uncontrolled diabetes | 5 months |
| 6 | I | IIIC | Renal failure | 6 months |
| 7 | IIIB | IIIC | Inadequate amphotericin | 6 months |
| 8 | VA | VA | Active Covid infection during first surgery | 3 months |
| 9 | VA | VA | Amphotericin intolerance | 1 month |

### Mortality

Among the cohort of 193 patients n=12 (6.2%) patients died. Among them n=5 patients were primary cases and n=7 were revision cases. The cause of death is charted in Table 7.

**Table 7:**

| **PRIMARY CASES** | Stage of disease | Cause of death | Time interval from surgery |
| --- | --- | --- | --- |
| 1 | IIIC | Acute MI | 10 days |
| 2 | VB | Systemic fungal disease | 1 month |
| 3 | VB | Acute PE | 20 days |
| 4 | IIIC | Carotid blowout | 2 months |
| 5 | VB | Cardio-respiratory arrest | 15 days |
| **REVISION CASES** |  |  |  |
| 1 | VB | Frontal abscess/meningitis | 16 days |
| 2 | IVA | Acute PE | 2 months |
| 3 | VB | Pre-existing liver failure | 1 month |
| 4 | IIIB | RF | 2 months |
| 5 | IIIC | RF | 2 months |
| 6 | IIIC | Systemic fungal disease | 1 month |
| 7 | IIIC | Systemic fungal disease | 1 month |

Abbreviations: MI – Myocardial infarction, PE -Pulmonary Embolism, RF- Renal Failure

### Statistical analysis

The normality of the variables was assessed using Kolmogorov-Smirnov test. Quantitative variables have been expressed as mean and median (IQR). Qualitative variables have been summarized using proportions. The distribution of the diseases in various sites between primary and revision cases was compared and the difference in proportions was statistically assessed using Chi-square test. Similarly, bilateral disease and orbital involvement between primary and revision cases were also assessed. A p-value of less than 0.05 was considered to be statistically significant. Analysis was done using SPSS software version 25.

## DISCUSSION

ROCM is an invasive, life-threatening opportunistic infection in immunocompromised patients. During the second wave, there was a steep rise in the number of mucor cases in India. Conditions such as hyperglycaemia, hypoxia, reduced phagocytosis, high ferritin levels, metabolic acidosis caused severe immunosuppression in post-covid patients. Write the study name here and say, As per the article written by Sannathimmappa MB et al, excess steroid administration for the treatment of covid infection in the presence of uncontrolled diabetes facilitated germination of fungal spores ^(3)^

## PATTERN OF SPREAD

The exact pathogenesis of rhino-cerebral mucormycosis and its locoregional spread are not clearly understood. It is generally believed that Mucorales infection results from inhalation of fungal spores which are normal commensals of the airway^(4-6)^ Spores can inoculate the nasal mucosa, spreading to the paranasal sinuses, orbit and finally the intracranial fossa, when the host immunity is compromised. ^(5, 6)^.

Mucorales are vaso tropic and are notorious for their ability to cause extensive vessel thrombosis and tissue infarction^(6, 7)^ The damage and penetration through endothelial cells lining the blood vessels is a critical step for dissemination.^(7, 8)^ In normal hosts, the spores produce a phagocytic response which prevents active infection. In diabetics and those with impaired immune systems, the fungal inhibitory responses are suboptimal, and germination ensues.^(9-11)^

In studies (Spellberg B etal and Ibrahim AS et al) it was observed that the presence of nonviable R.oryzae can lead to endothelial damage, in part explaining the inefficiency of antifungal drugs and surgical debridement as the mainstay of treatment^(7, 8)^. The eradication is difficult due to the aimless cycle of arterial invasion and occlusion, tissue hypoxia and necrosis, local acidosis, and thus, further extension of the fungal hyphae. In addition to tissue hypoxia, the arterial occlusion caused by this disease also makes the delivery of intravenous drugs to the site of infection difficult.^(8, 11)^

It was noted that the perivascular channels play a crucial role in dissemination beyond normal sinus walls. The peri-sinus spread is characteristic and may also involve the pterygopalatine fossa and infratemporal fossa and is believed to be along the neurovascular bundles.^(6)^ The infection spreads more along the vascular channels and nerves, rather than a contiguous spread of the tumor via the posterior wall of the maxillary sinus. There are several orifices that connect this space to others such as the inferior orbital fissure to the orbit, the pterygomaxillary fissure to the infratemporal fossa and the masticator space, the sphenopalatine foramen to the nasal cavity, the foramen rotundum and pterygoid canal to the middle cranial fossa and the greater palatine canal to the palate.^(6)^ Inferior orbital nerve involvement can explain the initial pain and paresthesia in this region.^(6)^ The mucor then spreads along the nerve to the facial soft tissues with an intact anterolateral wall.

Potential extra-arterial routes of orbital spread in mucormycosis include direct spread from the orbit and along infraorbital nerves. It may also result from spread through the nasolacrimal duct and the paper thin medial orbital wall. Vascular routes of ocular invasion include retrograde propagation from the cavernous sinus or ophthalmic veins or the embolization via the ophthalmic, central retinal, and ciliary arteries. Sclera is a natural barrier that inhibits direct fungal invasion except at the sites at which blood vessels and nerves pierce it, which represent areas of potential weaknesses. Thus, ischemia may result from occlusion of the internal carotid artery, ophthalmic artery, central retinal artery, or posterior ciliary artery. The internal carotid artery is one of the most frequently thrombosed vessels and is often found to be invaded.^(12^)

The primary ROCM cases demonstrated significant signs of early osteomyelitis in the pterygoid wedge and floor of the sphenoid sinus. The authors firmly believe that the posterior spread of the disease starts with the nidus in the pterygoid wedge and spreads through vascular channels in the bone towards the central skull base. The spread from the pterygoid wedge can also occur by venous sinusoids and Haversian canals which represent a good medium for harboring fungi. Fungus becomes established in the marrow and provokes thrombophlebitis leading to bone sequestration, which assists in the proliferation of hyphae, and produces an area of low oxygen tension. The impaired blood supply in the affected area causes sequestrum. The clival bone demonstrated maximum involvement due to high density of venous sinusoids.

The extension to the frontal sinus was noted from orbit, septum and ethmoidal sites. However, direct erosion of the frontal bone leads to subperiosteal abscess, subdural collections, meningitis, and encephalitis. The direct spread through the cribriform plate into the anterior cranial fossa may represent perineural spread via olfactory nerves.^(13^) Intracranial involvement can occur through the superior orbital fissure, ophthalmic vessels, perineural route, cribriform plate or the carotid artery.^(14)^

Due to an unexpected spike in the number of cases, increased patient load, lack of human resources, inadequate essential supplies, unavailability of universal guidelines and paucity of infrastructure, we experienced an initial struggle in the management of ROCM at our centre. Therefore, the need for an effective and systematic surgical staging system was realised and implemented with the help of a multidisciplinary team. The surgical treatment for mucormycosis over time is ‘debridement till healthy bleeding.^(15)^ However, we stress on ‘defining the debridement with anatomical boundaries’ for better surgical outcome. Unlike malignancies, the disease pattern in ROCM may not always be contiguous. Therefore, a higher stage does not necessarily mean involvement of structures in the lower stages. ^(15^)

Our study proposes a detailed protocol for the surgical management of invasive skull base fungal osteomyelitis, based on the radiological assessment of disease and its progression. This was done with the aid of various radiological imaging studies (MRI, CBCT, MDCT). After thorough analysis, a novel staging system was designed to standardize the reporting and management. This could facilitate the surgeon in counselling the patients scientifically, regarding the extent of involvement, probable surgical outcome and post-operative rehabilitation.

In the proposed staging system entire region of sinuses to skull base is divided into anterior and posterior compartments.

The anterior compartment is subdivided into rhino-palatal and rhino-orbital, which forms stage I and II of our classification respectively. The rhino-palatal stage is again subdivided into stage Ia and Ib.

Ia includes the premaxillary region and paranasal sinuses. This is approached via EMDA which gives access to pterygopalatine and infratemporal fossa. An additional trans-septal window is made for ‘cross-court’ access. The gross pathology varies from fat necrosis, thrombosis of internal maxillary artery, necrosis of pterygoid muscles and involvement of walls of maxilla. Once disease is cleared, pterygoid wedge is inspected (Fig5a,5b).

**Fig 5.**
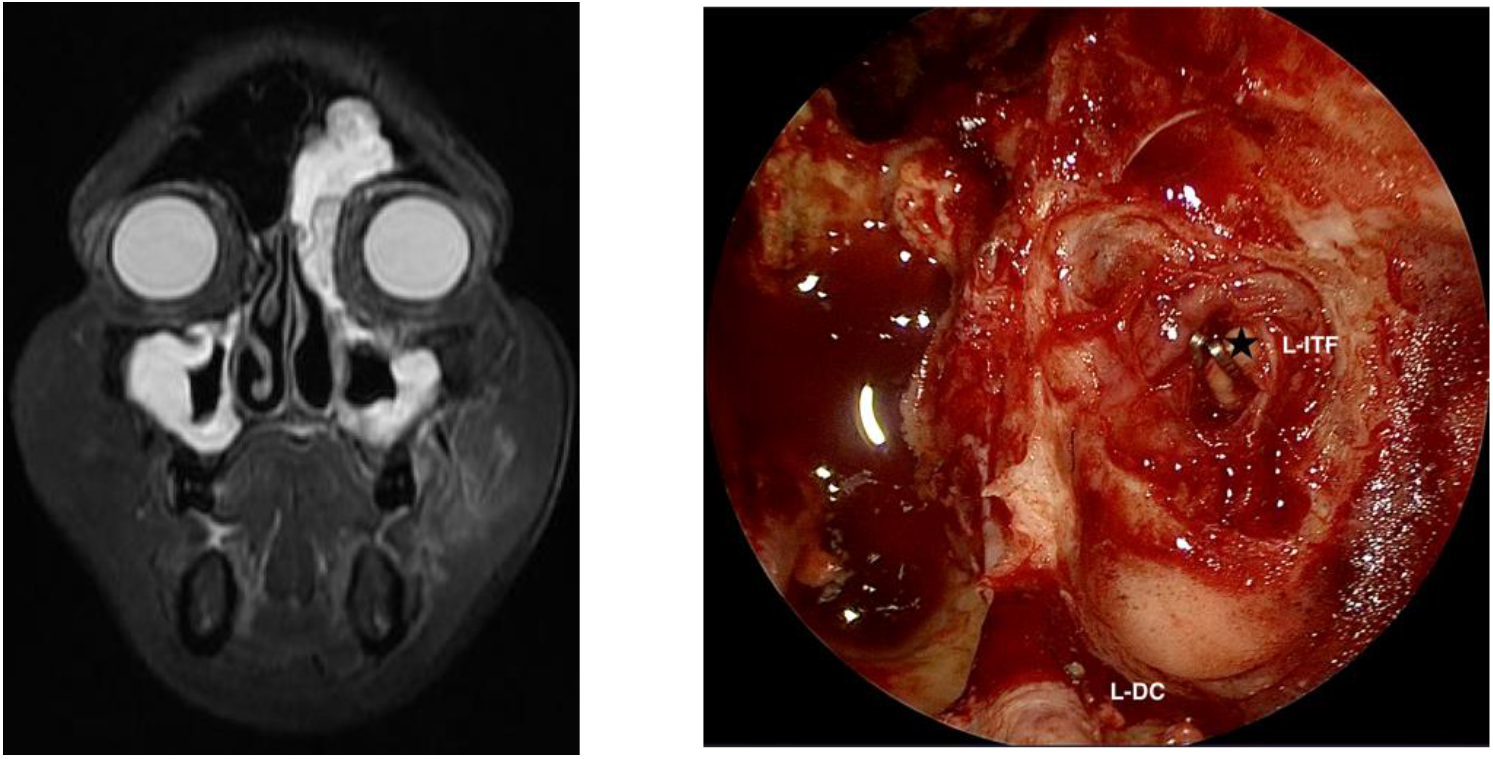
(a, b)

Stage Ib patients were managed with sub-labial segmental resection of palate with preservation of palatal mucosa with the assistance of facio-maxillary surgeon.

Stage II of the classification is divided into 4 sub-groups. Stage IIa includes involvement of the medial and inferior orbital walls, which is managed by endoscopic orbital decompression of medial and inferior wall. Stage IIb incorporates involvement of extraocular muscles and is managed by endoscopic trans-nasal trans-orbital extraconal globe sparing debridement.

Endoscopic trans-pterygoid exposure with clearance of superior orbital fissure is the management of patients in Stage IIc. Stage IId in which anterior and posterior globe is involved, the treatment of choice is orbital exenteration (Fig 6a, 6b). Oculoplastic surgeon administers Transcutaneous Amphotericin B (TRAMB) injection to all patients in stages IIa to IIc, at a dose of one ml of 3.5mg/ml of Amphotericin for 3 days.

**Fig 6.**
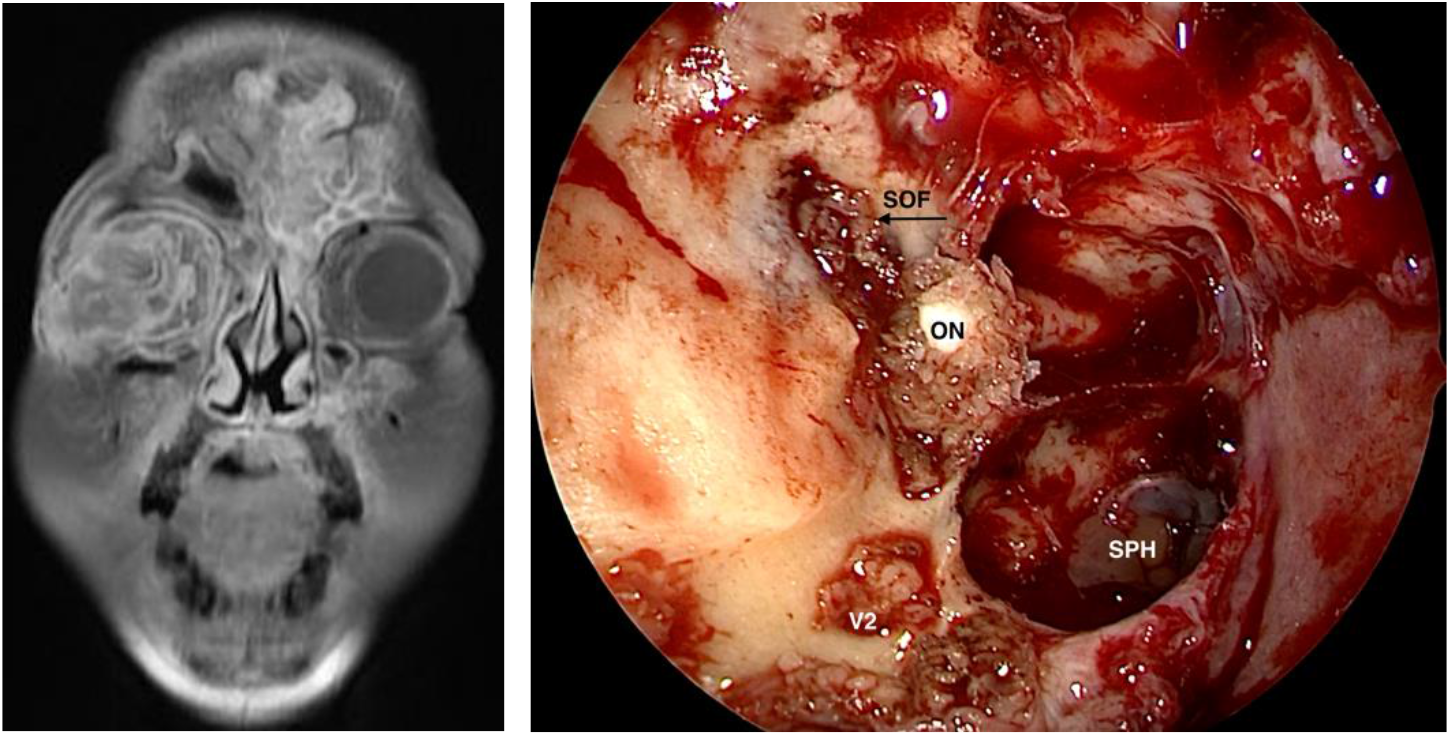
(a, b)

From stage III, posterior compartment with central skull base osteomyelitis (CSBO) commences. It is subdivided into A, B, C and D. Endoscopic midline trans-sphenoidal approach to clear the Sella, clivus, floor and lateral recess of sphenoid summarises the management of Stage IIIa. Clearance for stage IIIb requires a trans-pterygoid supra-petrous approach to Meckel’s cave and cavernous sinus. Similarly, Stage IIIc also requires a trans-pterygoid approach and supra-petrous approach to base of temporal lobe (Fig 7a,7b). In stage III disease we noticed that those cases with involvement of clivus, floor of sphenoid sinus and bilateral greater wing of sphenoid, following complete surgical clearance gave the appearance of a ‘cross’ sign in the post-operative MRI. We have termed this as ‘Holy-Cross Sign’ (Fig 8). Stage IIId is frank palatal disease with mucosal involvement for which trans-facial approach and maxillectomy is done.

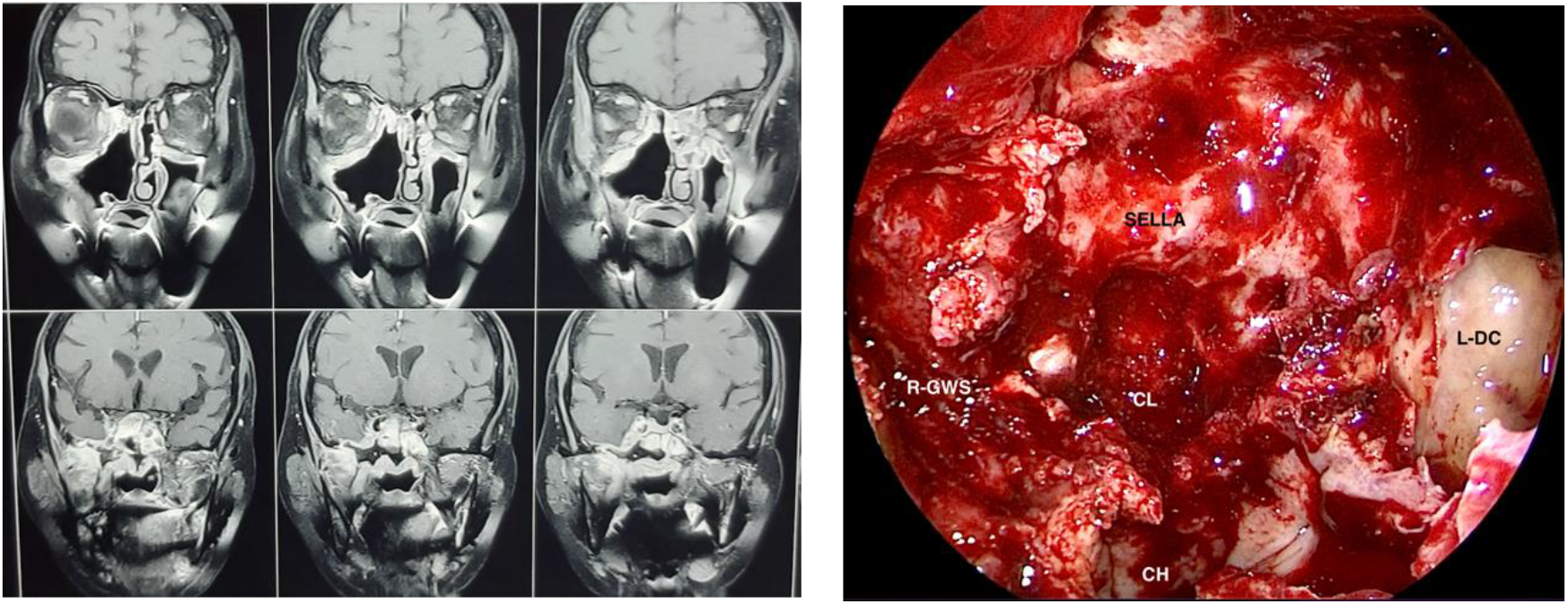

**Fig 8.**
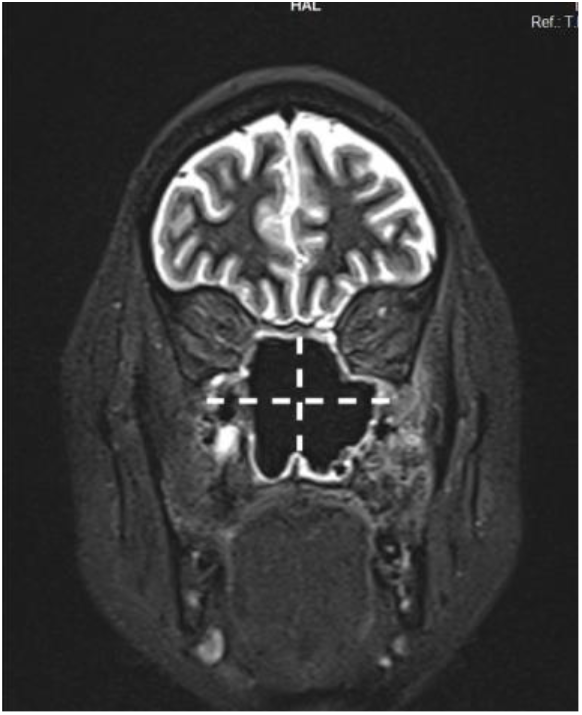

Involvement of bones of the cranium viz., frontal, parietal, temporal, zygoma and superior orbital wall is managed via external approaches in combination with endoscopic approach (Fig 9a, b, c).

**Fig. 9.**
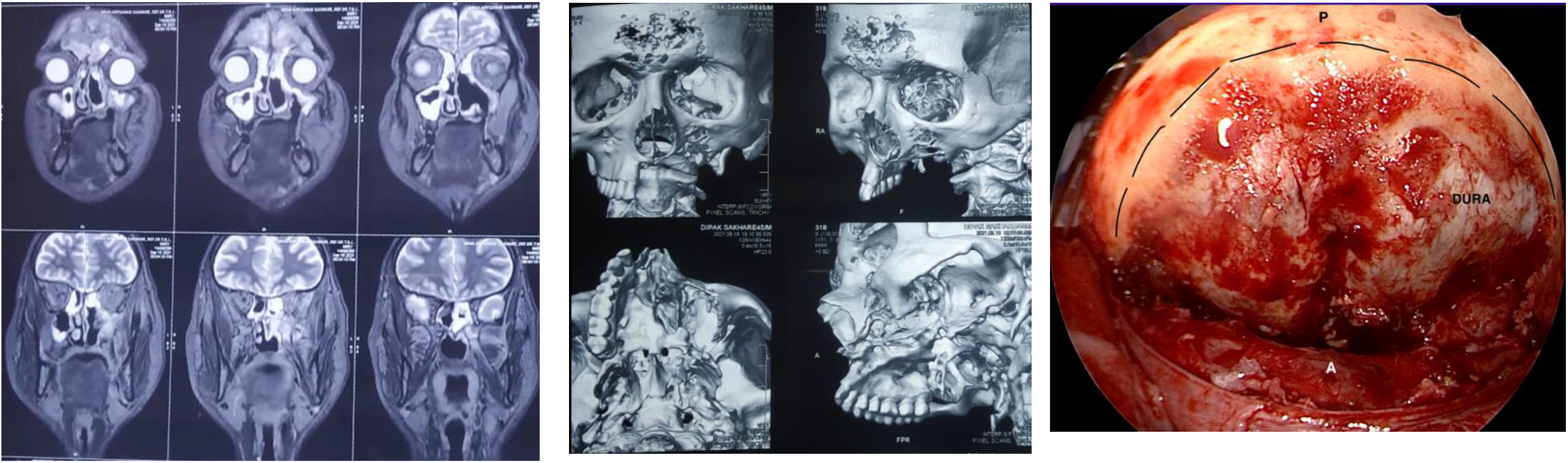
(a, b, c)

**Fig 10.**
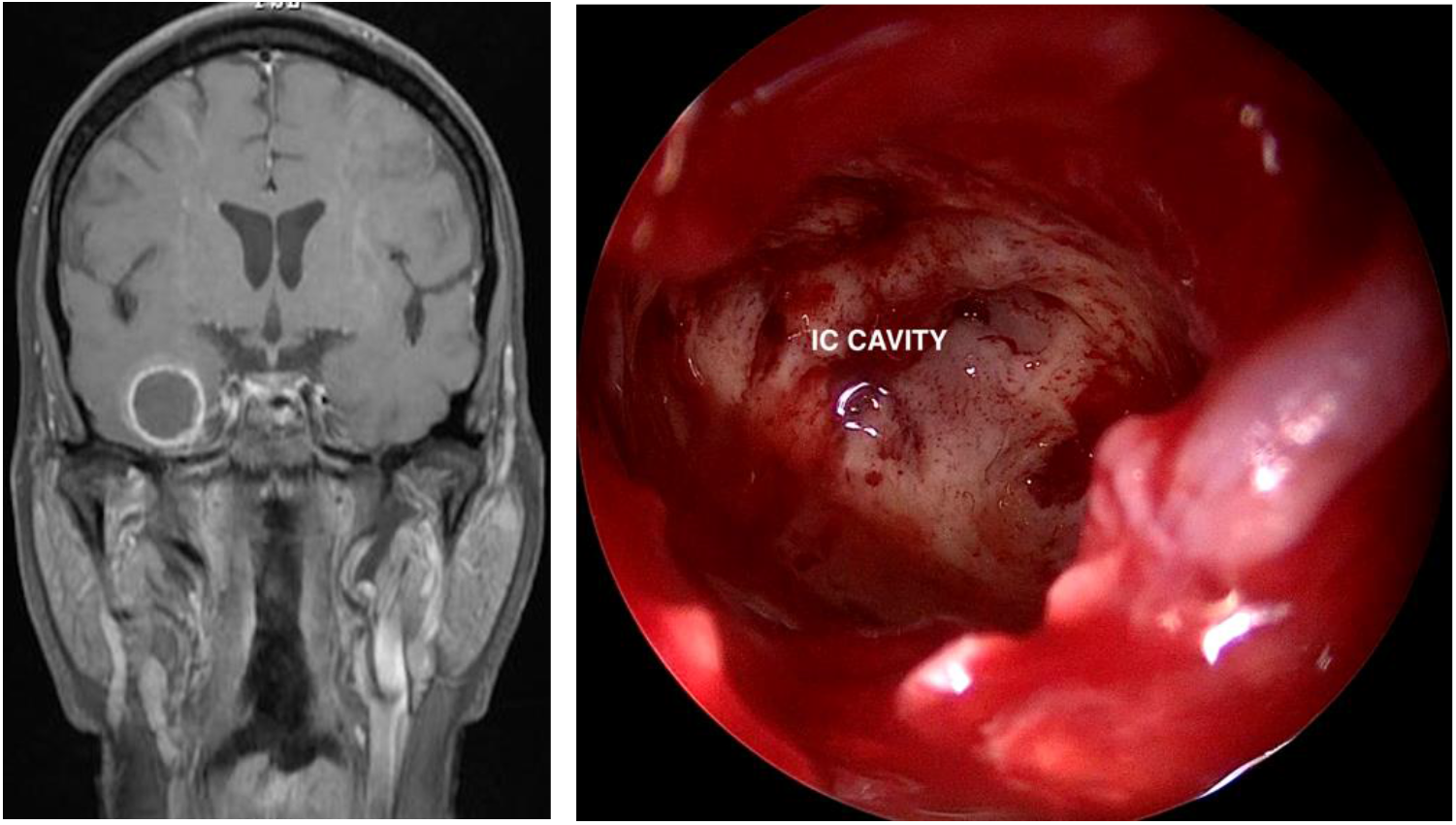
(a, b)

Intracranial abscess in frontal and temporal regions may be drained endoscopically (Fig 9a,9b) whereas other areas might need a combination with trans-cranial approaches.

An extensive review of literature revealed multiple staging systems proposed by various authors; Honavar SG et al (n=2669) described a 4 tier staging based on anatomical progression of disease, Soni K et al (n=145) proposed another 4 tier staging based on clinical and radiological evaluation, Vaid N et al (n=65) described a severity grading system. A guideline for management for mucormycosis by Cornely OA et al was found, but this study was on the overall management rather than surgical protocol.^(16-19)^ The JSPM staging (n=193) in the current study is based on radiological imaging and has defined a debridement protocol based on anatomical landmarks.

Total number of patients in the cohort was 193 of which 79.8% were males and 20.2% were females. A male preponderance was seen in our study which was consistent with other studies in literature.^(19, 20)^ This could be due to other factors like increased smoking and compromised airway mucosa with poor inherent immune response in males, but this requires further studies. The mean age was 48.9 years. Young diabetic patients under 40 years consisted of 42% of the total cohort, indicating the rising trend of diabetes in the younger Indian population.

A total of 129 cases (66.8%) accounted for primary and 64 (33.2%) for revision cases. Due to the lack of amphotericin and delay at presentation, two-third of revision patients presented with advanced stages (60.9% in stage IIIc and IVa) requiring central skull base clearance with combined approaches.

According to a study conducted by Michael Dan, the spectrum of head and neck Mucormycosis consists of 50% of the total presentations ^(21^). The spectrum of disease patterns described in this study are; Rhino-palatal, Rhino-Orbital, central skull base osteomyelitis, cranial bone involvement and intracranial disease.

Our patient cohort consisted only of post-covid ROCM patients with comorbidities. Out of all patients 9.4% of patients did not have any of the traditional risk factors discussed above. In a study by Patel et al and another study by Soni K et al they reported 12% and 11% of total patients of ROCM without any co-morbidities respectively.^(16, 22)^ Therefore, an afterthought, whether the disease is due to the viral infection or due the treatment for it, is not clear.

It is a well-established fact that diabetes is the most common predisposing factor for Mucormycosis. Injudicious use of corticosteroids especially in diabetic patients causes severe hyperglycaemia, ketoacidosis and reduced phagocytosis further impeding the host immunity. Among our total patients 90.6% of patients were diabetics and 78.7% were treated with corticosteroids.

‘Global guidelines for the management of mucormycosis (2019)’, a study by Cornely OA et al preferred MRI only in orbital and intracranial extension, whereas in our study MRI was the primary radiological investigation for all patients.^(17)^ In ROCM, the time interval between diagnosis and treatment is highly critical, because any delay in the treatment increases mortality by double fold^(23)^. CT scans tend to miss out on early diseases without bone erosion and thus increase this interval and accelerate the mortality rate. In ROCM, the following patterns may be seen in MRI; homogenous, heterogenous or non-enhancing in post contrast imaging and hypointense or hyperintense in T2W series. In Diffusion weighted imaging (DWI) lesions exhibit diffusion restriction. ^(24-26)^ CBCT was done in patients with palatal symptoms and in those where MRI showed alveolar/premaxillary involvement. MDCT was done in those patients with gross bone erosion.

Histopathological analysis of specimens from all cases confirmed the presence of pauciseptate fungal hyphae suggestive of mucormycosis.

Statistically significant number of cases with anterior disease had maxillary involvement and cases with posterior disease had involvement of pterygoid wedge, thus supporting our hypothesis of maxilla and pterygoid wedge being the epicentre in anterior and posterior disease respectively. Cutaneous involvement was seen in very few cases (n=4) in our study.

Figure 2 (Radiological staging) was compared to Figure 3 (Intra-operative staging) and it was noticed that when compared to MRI staging, in intra-op staging two patients down staged and three patients upstaged. Upstaging was possibly because the time period between MRI and surgery was more than two weeks, since neither of the patients agreed for surgery initially.

Therefore, by the time patients were taken up for surgery, the disease had progressed. In the other three patients who down-staged, MRI had over-diagnosed the inflammatory changes, but intraoperatively no disease could be appreciated. Thus, the importance of correlating radiological findings with intraoperative findings is extremely important.

Perineural spread via involvement of vidian nerve (37.8%), V2 branch of trigeminal nerve (39.5%), facial nerve (2.0%) and olfactory nerves (1.5%) were observed in our study. In a study by Parsi et al, perineural involvement of trigeminal root and middle cranial fossa via retrograde spread from infraorbital nerve and neurovascular bundle of pterygopalatine fossa has been described.^(27)^

Eighty two percent of our patients, at the time of presentation, presented with ROCM with involvement of the central skull base (36.5% primary and 43.7% revision cases). Sphenoid sinus, clivus, pterygoid wedge, greater wing of Sphenoid and posterior orbit were significantly more involved in revision cases. Paraclival internal carotid artery was involved in 31.2% and 33.3% of primary and revision posterior disease respectively.

Inadequate clearance of frontal sinus in primary disease results in spread to frontal tables and crista galli, thus supporting the significant increased percentage (37.1%) of cases in revision. Though temporal and frontal abscesses were also found more in revision patients (16.7% and 11.7% respectively), this finding was not statistically significant, probably due to the smaller number of cases. Almost all cases with involvement of zygoma and temporal fossa were seen among revision.

There are no definitive criteria to differentiate between residue and recurrence in Mucormycosis. In our study residual disease was seen in 4.6% of cases which is comparatively much less compared to other contemporary studies. ^(15)^

Out of the total cohort, 94.8% patients had no evidence of disease during follow-up, ranging from 6 to 18 months.

A total of 12 patients (6.2%) died. The mortality rate was also comparatively lesser when compared to the recent studies.^(15, 19)^

## SURGERY

Considering the abrupt dissemination of infection from the skull base to neurological structures and the poor availability of drugs in necrotic tissue, aggressive rhino-orbital and skull base debridement is advocated. In advanced cases, debridement of devitalized, infected and inflammatory tissue improves drug concentration, morbidity and mortality. However, limited availability of contemporary treatments burdens healthcare systems in low and middle-income settings. The primary trans-nasal access was through Endoscopic Modified Denker’s approach (EMDA), which aids in ‘four handed bi-nostril technique’. In this technique, the operating surgeon can use both his hands, instead of holding the endoscope in the left hand. The endoscope is held by an assistant, who can simultaneously do warm saline irrigations over the endoscope to prevent fogging. The scope is held at 11 O’ clock position utilizing the elasticity of the ala of the nose. This technique allows better visualization of the disease and the advantage of a panoramic view. EMDA also facilitates trans-pterygoid approaches and aids in post-operative inspection of operated areas to pick up early disease changes.

Surgical treatment of mucormycosis has been defined over time as ‘debridement until fresh bleeding’ ^(15)^. But we stress on ‘defining the debridement with anatomical boundaries’ for better surgical outcome.

## Conclusion

The second wave of covid-19 infection was accompanied by a sudden surge in the number of patients with ROCM. Multiple staging systems and treatment modalities have been described in the literature. Well defined guidelines of management of central skull base osteomyelitis has been unclear. Our new staging system can help surgeons on the step-by-step approach to central skull base debridement respecting the anatomical landmarks with minimal complications. The surgical treatment protocol proposed in this study helps in achieving better surgical outcomes with minimal residual disease.

## Data Availability

All data produced in the present study are available upon reasonable request to the authors

## SUMMARY

Rhino-orbito-cerebral mucormycosis presented to the Indian subcontinent as an endemic within a pandemic. This study was conducted in our tertiary care centre and a total of 193 patients were included. We studied our patient population systematically and analyzed the spread, progress and various clinical manifestations, and thus could arrive at a classification which could be used to prognosticate and form protocols in planning treatment strategies. Earlier classifications on this pathology did not stress on the importance of central skull base clearance for improving surgical outcome, whereas in our study two-thirds of patients underwent central skull base debridement. Endoscopic endonasal approaches were used in all cases in combination with external approaches, wherever it deemed necessary. We strongly believe that this article will open new vistas in the management of skull base mucormycosis.

